# Financial burden of cervical cancer in Bhutan and Zambia: a multiple low- and middle-income country cost survey

**DOI:** 10.1101/2025.11.05.25339642

**Authors:** Ahmad Fuady, Pempa Pempa, Charlotte Kasempa, Namakau Nyambe, Vanessa Tenet, Eric Lucas, Irene Man, Partha Basu, Iacopo Baussano

## Abstract

**Background:** Zambia and Bhutan have a high prevalence of cervical cancer (CC). The financial burden on affected households is substantial yet potentially mitigable through effective interventions. This study assessed the financial burden of CC in these countries by quantifying the proportion of households facing catastrophic costs and exploring their financial coping mechanisms.

**Methods:** Between January and July 2023, two cost surveys were conducted using the COEUS (Costing Surveys to Assess the Economic Burden of Cervical Cancer in Society) framework, from a societal perspective to assess household financial burden. In Bhutan, we used an outreach-based survey, while in Zambia, the surveys were completed at a health facility. Women or members of their household with CC were interviewed using a standardized questionnaire. We excluded individuals with multiple cancers or communication barriers. The survey captured direct medical, direct non-medical, and indirect costs related to diagnosis through treatment, including information on alternative remedy expenses. Catastrophic expenditure was defined as out-of-pocket expenses exceeding 10% of the annual household income. We also assessed perceptions of financial hardship according to households and coping strategies. All costs were reported in 2022 International Dollars (I$).

**Results:** Out of 164 interviews, 162 responses were analysed: 87 from Bhutan and 75 from Zambia. The total costs in Bhutan were higher than the spending in Zambia (median[min-max] I$4,025[90–4,0912] vs. I$1,703[315–11,059]). This was mainly due to treatment costs at professional health care facilities, and significantly more in Bhutan than Zambia for alternative remedies (I$587[0–15,749] vs. I$0[0–3,036]). In Bhutan, higher costs were reported among households of women with stage 2 CC, while in Zambia, higher costs were reported among households of women with distant cancer. Over half of affected households lost income, and the majority faced catastrophic costs: 85% in Bhutan and 89% in Zambia. Most Zambian women (68%) reported that cervical cancer severely impacted their financial situation and coped by seeking help from relatives, religious bodies, or non-government organizations (72%) as well as reducing food and other purchases (69%).

**Conclusion:** CC care seeking and treatment often have catastrophic financial consequences to affected women living in low- and middle-income countries. Strengthening CC prevention through HPV vaccine, screening, and early detection are required to reduce the incidence of catastrophic costs due to CC.

## Background

Cervical cancer (CC) is the fourth most common cancer among women worldwide, with an estimated 662,301 new cases and 348,874 deaths in 2022.[1] The largest proportion of CC is in low- and middle-income countries (LMICs) and in many countries of sub-Saharan Africa CC remains the most common cancer among women.[2, 3] Despite substantial resource limitations, an increasing number of LMICs are adopting key public health measures to attain cervical cancer elimination.[4] Two LMICs that have prioritized cervical cancer prevention are Zambia and Bhutan.

Zambia has one of the highest rates of CC worldwide, with an age-standardized annual incidence and mortality of 71.5 and 49.4 per 100,000 women in 2022, respectively. To tackle this problem, Zambia introduced Human Papillomavirus (HPV) vaccination in 2019[5] and provides free screening to women aged 30-59 years through the Cervical Cancer Prevention Program in Zambia,[6] with HPV-DNA testing under a pilot programme that was initiated in 2021.[7] These initiatives aim to align Zambia with the WHO’s global Cervical Cancer Elimination Initiative goals for 2030.[2, 7] Another LMIC which successfully introduced HPV vaccination and cervical cancer screening is Bhutan. Bhutan’s national HPV vaccination program was launched in 2010 and has consistently high coverage around 90%.[8] HPV-DNA-based CC screening was successfully introduced nationwide in 2019.[9–11]

In both countries however, despite political commitment from local stakeholders to screening and vaccination programs, women with CC are still facing structural challenges, particularly for diagnosis and treatment, such as limited infrastructure and limited access to healthcare facilities for some subgroups of women. These challenges may be exacerbated by the financial burden of CC management on women, and their households. Women with CC need access to expensive CC-related services, such as surgery, radiotherapy, brachytherapy, and chemotherapy, which have been shown to cost as much as $71,000 in some LMICs.[12] Furthermore, although some countries, like Bhutan, offer free universal health coverage, women being screened or diagnosed with CC, as well as their households, may still encounter substantial direct non-medical costs (e.g., transportation, food, and accommodation costs) and indirect costs (e.g., income loss) [12]. Additionally, this cancer predominantly affects women at a young age, imposing significant productivity losses due to disability and a substantial loss of life-years resulting from premature death.[13, 14] All of these factors may contribute to an elevated financial burden on households,[15–17] which can be beyond their capacity, leading to ‘catastrophic’ costs—defined as health-related spending exceeding 10% of the annual household income.

Very few data about the cost of CC screening and management from the societal perspective are available from LMICs. To start filling this gap, this study assessed the financial burden of CC among households in Bhutan and Zambia, by estimating the out-of-pocket costs, quantifying the proportion of households experiencing catastrophic costs, and exploring how households perceive and cope with this cost burden.

## Methods

### Study design

We conducted two cross-sectional surveys: one in Bhutan, the other in Zambia, between January and July 2023 to assess the financial burden of CC on women and their households, in both countries. We have previously developed a master study protocol, based the COEUS (Costing Surveys to Assess the Economic Burden of Cervical Cancer in Society) framework for costing surveys, which assesses financial burden from a societal perspective.[12] We adapted the protocol to the local health system setting of each country, and then decided to follow two different approaches based on practical feasibility. In Bhutan, we adopted an outreach-based cost survey approach, while in Zambia, we used a facility-based cost survey approach.

### Study participants and recruitment

We aimed to establish nationwide representative estimates for both countries. In Bhutan, potential participants were identified from national cancer registries based on available contact information. We included women lost to follow-up and relatives of deceased women with CC. Participants were selected retrospectively, starting from the most recent CC cases and stratified by cancer stage (1-4 or local to distant). If a woman appeared multiple times in the registry, her latest diagnosis determined inclusion. When first contacted, interviewers applied exclusion criteria. With consent, interviews were scheduled either face-to-face or by phone, based on feasibility. All interviews were conducted by trained interviewers.

In Zambia, a facility-based cost survey was conducted at the Cancer Diseases Hospital (CDH), the national referral centre for CC. Women with CC attending scheduled gynaecologic visits at the clinic were consecutively recruited, as outlined in the COEUS framework. Recruitment continued until the required sample size was reached.

In both countries, we excluded women who were diagnosed with multiple cancers and/or with mental illness affecting their ability to engage in the interview, provide a reliable response, or who could not be contacted and/or could not communicate clearly or be assisted, if necessary, by a family member(s) during the interview.

### Sample Size

As there were no available data on catastrophic costs due to CC for both countries, we based our sample size on an Indian study, where 62% of CC-affected households spent more than 10% of their annual household income.[15] Using this estimate, with α=0.05 and β=0.1, a minimum of 91 participants per country was required. We aimed for equal distribution across cancer stages. In Bhutan, we used the national cancer registry to allocate 25 participants to each stage (1-4), targeting 100 women. Households of deceased women diagnosed with CC were also included. Staging followed local classification (local, regional, distant), and 75 women attending the Cancer Diseases Hospital were targeted for recruitment. Households were approached consecutively until the sample size was met. (Figure 1)

**Figure 1.**
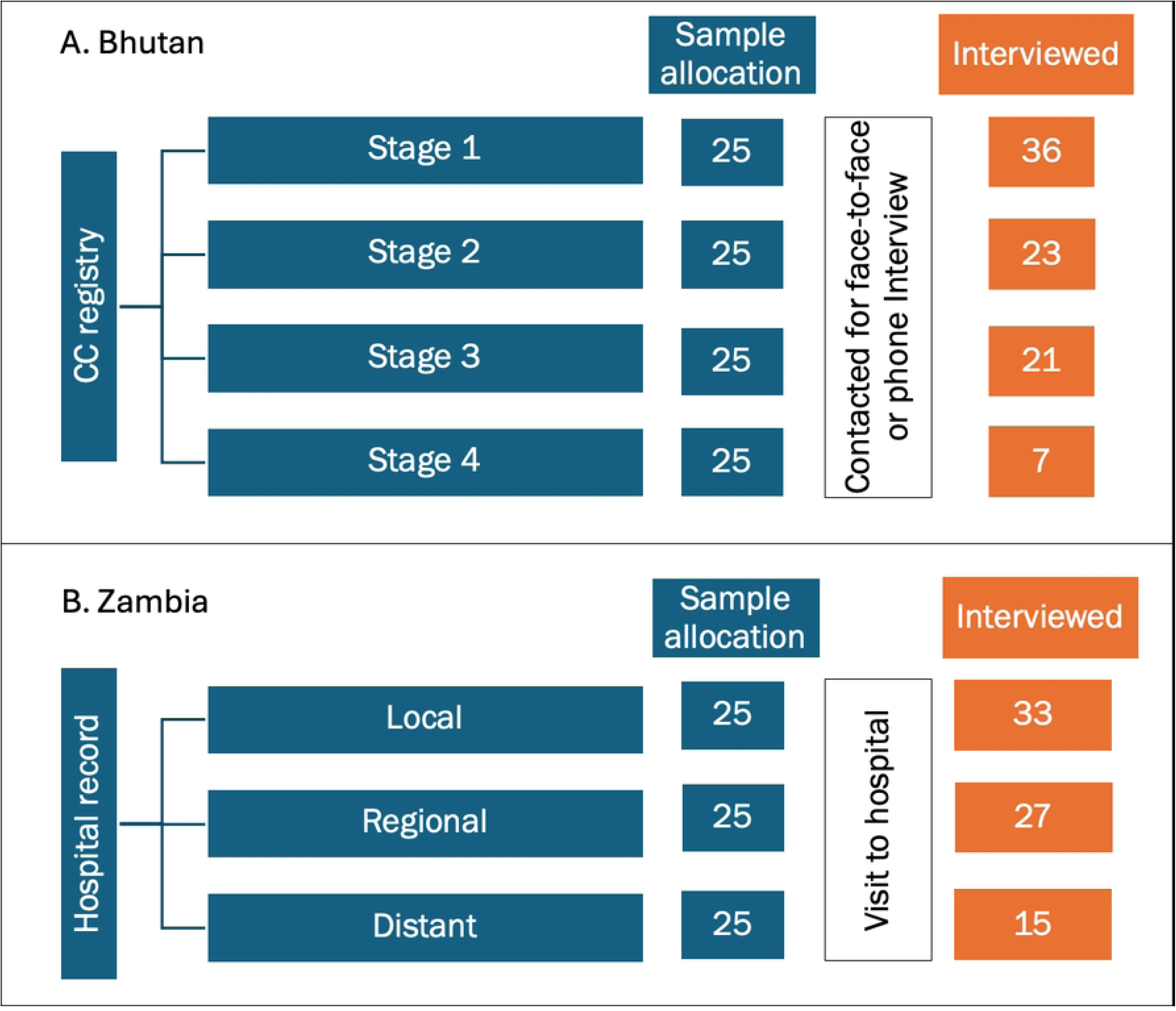
Sample size allocation and selection, *CC=cervical cancer*

### Instruments and Interviews

Before recruitment, we developed a questionnaire based on previous costing surveys on tuberculosis and similar studies.[15, 18] Local experts in Bhutan and Zambia reviewed the content to ensure cultural and health system relevance. The questionnaire included both quantitative and qualitative questions, organized into six sections: (1) personal identification, (2) demographics, (3) pre-diagnostic and diagnostic costs (including screening), (4) treatment costs, and (5) perceived financial burden of CC.

In Bhutan, interviews were conducted by healthcare workers familiar with CC terminology to ensure accurate reporting of costs. In Zambia, the interviews were conducted by doctors and nurses at the CDH. All interviewers were trained how to conduct the interviews. While the questionnaire was written in English, interviewers were allowed to conduct interviews in Dzongkha (Bhutan), Nyanja and Bemba (Zambia), or other local dialects, with help from a translator when needed. All responses were first recorded on paper, and then entered on the Redcap platform for data management.

### Variables Costs

Costs were assessed during two main phases: (a) pre-diagnostic and diagnostic and (b) treatment phases. The pre-diagnostic and diagnostic phase covered the period from symptom onset to diagnosis, including costs related to clinical examinations and screening, if applicable. The treatment phase included costs from four types of visits: (a) for pre-cancerous lesions (e.g., cryotherapy, loop electrosurgical excision procedure (LEEP), thermal ablation), (b) for cancerous lesions (e.g., surgery, radiotherapy, chemotherapy), (c) for non-formal care, such as traditional, religious, or alternative remedies including any supplements, and (d) for follow-up visits after completing specific treatment cycles.

Costs in each phase were categorized as direct medical costs (DMCs), direct nonmedical costs (DNMCs), and indirect costs (ICs). DMCs included administrative fees, treatments (including traditional remedies), and examination fees. DNMCs included fees for registration, examinations, treatments, and traditional remedies. DNMCs included travel, accommodation, and food expenses for the woman with CC and accompanying household members. ICs referred to income loss due to hospital visits or hospitalization, calculated based on self-reported expected losses.

In Bhutan, where public healthcare services are free, DMCs at public facilities were recorded as zero. The government also provides daily allowances for women referred abroad for treatment (e.g., in Thailand or India); DNMCs were adjusted accordingly based on reported allowances. Total costs were calculated as the sum of DMCs, DNMCs, and ICs for both phases.

### Household income

Household income included all earnings from income-generating household members, such as formal and informal wages, sales (e.g., agricultural products), pensions, rentals, inheritance, donations, and government or non-government organizations (NGOs) support (e.g., cash transfers). For income-earning women, we asked whether their wages were received daily, weekly, monthly, or irregularly (e.g., seasonal farming). If income from a family business could not be broken down by family member, the amount was recorded as combined household income. We asked about income before CC diagnosis and calculated annual household income for catastrophic cost analysis.

Recognizing potential difficulties in accurately reporting income and expenses, we also collected qualitative data to better understand participant responses. Some participants reported only total estimates rather than itemized costs, while others had irregular income sources. These qualitative insights supported the analysis and helped to validate responses.

Participants reported costs and income in local currencies: Ngultrum (Nu) in Bhutan and Zambian Kwacha (ZMK) in Zambia. We adjusted for inflation to 2021 local currency values and converted the amounts to 2022 International Dollars (I$) using World Bank exchange rates.

### Financial burden

We assessed the financial burden of CC by assessing: 1, household income reduction after diagnosis, 2, perceived financial burden (no, little, moderate, or severe impact), 3, coping mechanisms, and 4, catastrophic costs. Definitions of catastrophic costs varied;[20, 21] in this study, costs were considered catastrophic if total expenses exceeded 10% of a household’s annual income. Sensitivity analyses were conducted using more conservative thresholds of 15%, 20%, 25%, and 30% to evaluate the robustness of our findings.

## Results

We interviewed 89 participants in Bhutan and 75 participants in Zambia. Two Bhutanese responses were excluded because of incomplete cost data, leaving 87 from Bhutan and 75 from Zambian for analysis. In Bhutan, CC stages were stage 1 (41%), stage 2 (26%), stage 3 (24%), and stage 4 (8%). In Zambia, stages were local (44%), regional (36%), and distant (20%) cancer.

The mean age of participants was similar: 54 years in Bhutan and 53 in Zambia. Most Bhutanese participants were married (77%) and had no formal education (79%). In Zambia, 55% had health insurance and 81% received financial support from the government or NGOs. In both countries, the majority of women with CC were income earners. (**Table 1**)

**Table 1.**
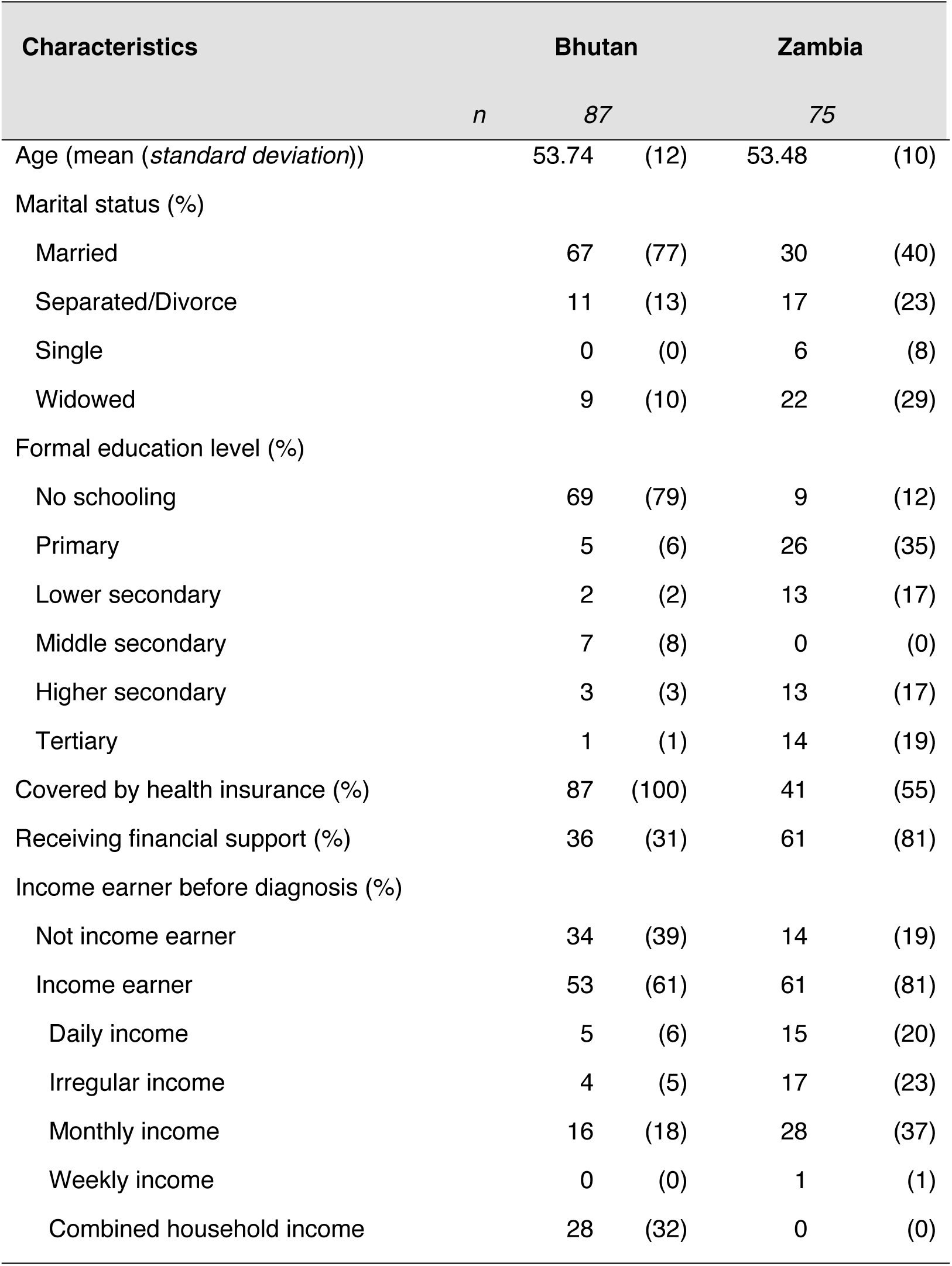
Participant characteristics.

CC-related costs were differently distributed in both settings and higher in Bhutan than in Zambia. (**Table 2**) Most costs in both countries occurred during the treatment phase in formal healthcare facilities. DMCs were higher in Bhutan, while Zambian participants reported higher DNMCs, such as transportation and food. Cost variation was also greater in Bhutan, with a wider range of spending across districts compared to Zambia. (See **Supplementary Table 1**)

**Table 2.** Cervical cancer-related costs in Bhutan, in I$.

| Costs | Bhutan |  |  | Zambia |  |  |
| --- | --- | --- | --- | --- | --- | --- |
|  | Median | Min | Max | Median | Min | Max |
| Prediagnostic and diagnostic costs |  |  |  |  |  |  |
| Direct medical costs | 0 | 0 | 226 | 39 | 0 | 906 |
| Direct non-medical costs | 63 | 0 | 4,440 | 56 | 0 | 995 |
| Indirect costs | 0 | 0 | 1,020 | 0 | 0 | 750 |
| Total | 80 | 0 | 4,440 | 134 | 0 | 1,838 |
| Treatment at formal healthcare |  |  |  |  |  |  |
| Direct medical costs | 0 | 0 | 36,101 | 281 | 0 | 2,155 |
| Direct non-medical costs | 745 | 0 | 14,260 | 551 | 64 | 3,467 |
| Indirect costs | 0 | 0 | 15,343 | 0 | 0 | 8,746 |
| Total | 1,814 | 0 | 36,101 | 1,149 | 125 | 10,614 |
| Alternative/traditional remedies |  |  |  |  |  |  |
| Direct “medical” costs | 451 | 0 | 15,343 | 0 | 0 | 1,012 |
| Direct non-medical costs | 0 | 0 | 406 | 0 | 0 | 2,024 |
| Indirect costs | 0 | 0 | 677 | 0 | 0 | 984 |
| Total | 587 | 0 | 15,749 | 0 | 0 | 3,036 |
| Follow-up visits |  |  |  |  |  |  |
| Direct medical costs | 0 | 0 | 4,061 | 0 | 0 | 876 |
| Direct non-medical costs | 59 | 0 | 8,123 | 307 | 9 | 2,062 |
| Indirect costs | 0 | 0 | 812 | 0 | 0 | 781 |
| Total | 140 | 0 | 8,123 | 411 | 9 | 2,062 |
| Total costs |  |  |  |  |  |  |
| Direct medical costs | 1,083 | 0 | 36,327 | 337 | 4 | 2,374 |
| Direct non-medical costs | 1,101 | 0 | 14,260 | 1,061 | 159 | 4,334 |
| Indirect costs | 0 | 0 | 15,433 | 0 | 0 | 8,933 |
| Total | 4,025 | 90 | 40,912 | 1,703 | 315 | 11,060 |

High and varied total costs in Bhutan were particularly apparent among women with stage 2 CC, while the costs among women with stage 1 and stage 3 CC—despite the wide range—were generally below I$10,000. (**Figure 2**) Zambia had a different distribution of costs per stage. Few women with local CC incurred more than I$5000, and women with distant CC incurred higher expenditure compared to those affected by local and regional CC.

**Figure 2.**
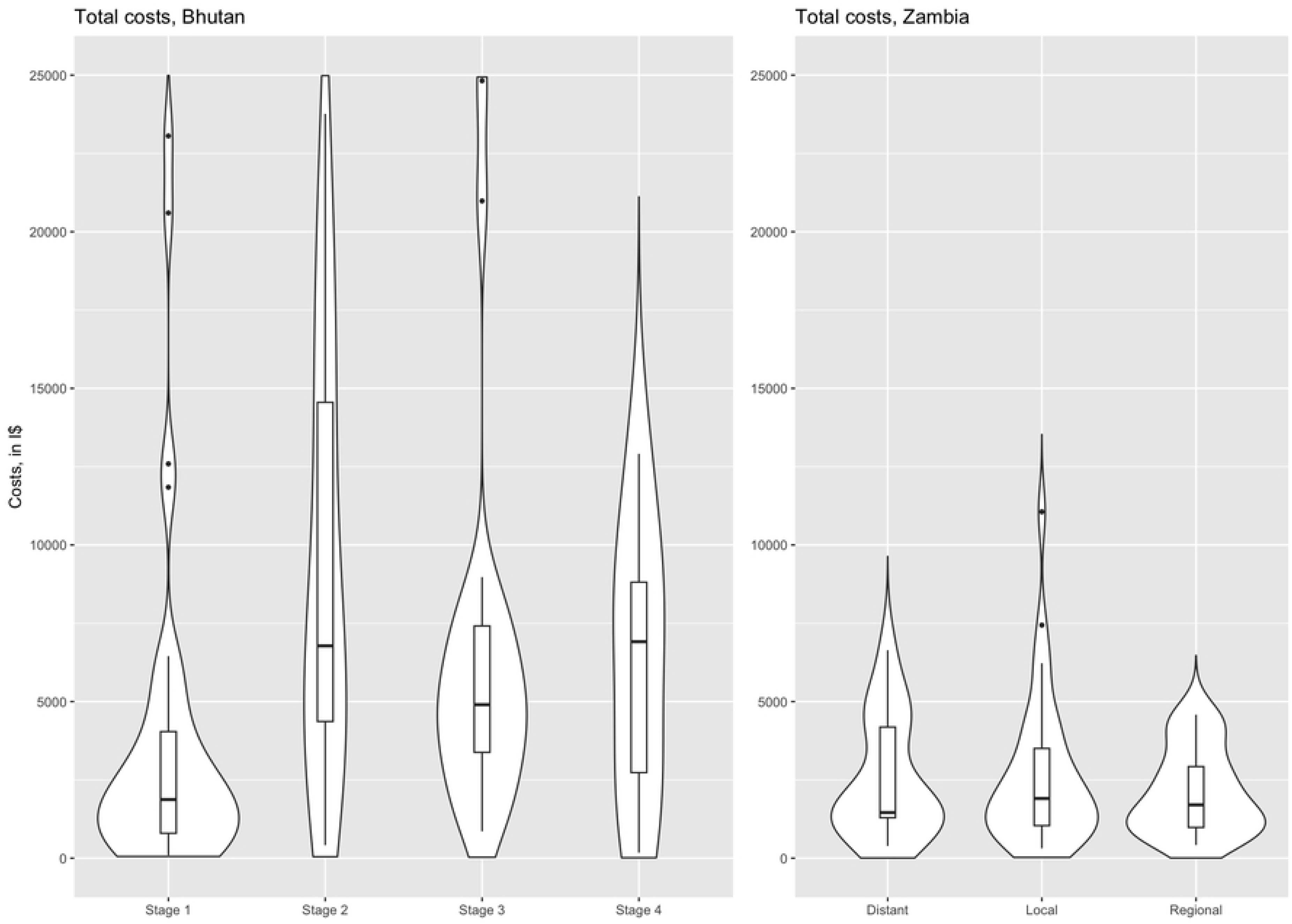
Cervical cancer-related cost by stage at diagnosis.

There was a higher number of women accessing alternative or traditional medicine in Bhutan than in Zambia. (**Table 3**) About 75% of women with CC in Bhutan sought non-formal healthcare remedies, such as Lama—someone who practices traditional healer in Bhutan, at different stages: stage 1 (72%), stage 2 (87%), stage 3 (71%), and stage 4 (57%). In Zambia, there was no common religious remedy reported, but 27% of women diagnosed with distant CC visited traditional healers. The higher frequency of visits to alternative remedy providers among Bhutanese women with CC resulted in higher costs than those among Zambian women. (See **Supplementary Figure 1**)

**Table 3.**
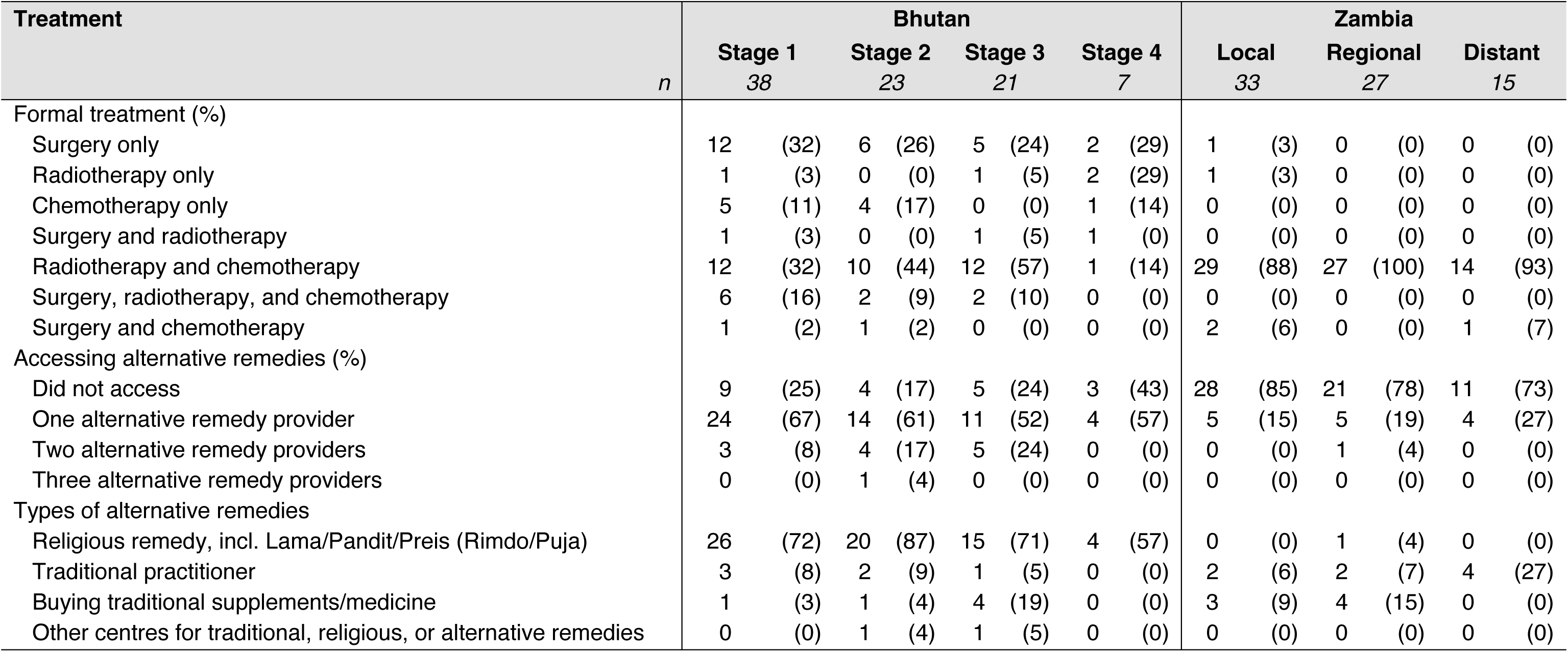
Treatment modalities accessed by women with cervical cancer in Bhutan and Zambia, according to stage.

This survey included comment boxes for interviewers to complement quantitative cost data with qualitative insights. These explanations helped clarify unreported costs, for example, *“(Two) patient(s) were referred from a regional hospital to a national referral hospital in Thimphu using ambulance*”, resulting in no reported transportation costs. Other comments examples are: “*the patient was a staff of the regional hospital and resides in the hospital compound, hence no cost in travel, food and accommodation*” and *“ritual (alternative remedies) was conducted by spouse”*, leading to no reported DMCs.

Not all participants provided household income data: 15 participants in Bhutan and 10 participants in Zambia left income questions unanswered. Among participants who reported their income, over half reported a loss in income. (**Table 4**) In Zambia, a severe financial impact was perceived by 68% of participants. To cope with financial challenges, 72% sought help from relatives, religious bodies, or NGOs, and 69% reduced food and other purchases. In Bhutan, moderate to severe financial burden was perceived by 43% of participants, and 74% relied on personal savings.

**Table 4.** Financial burden faced by cervical cancer-affected households.

| Financial burden | Bhutan |  | Zambia |  |
| --- | --- | --- | --- | --- |
| Income loss (%)* | 40/53 | (75) | 34/61 | (56) |
|  | N=74 |  | N=65 |  |
| Perceived financial burden (%) |  |  |  |  |
| No impact | 25 | (34) | 1 | (2) |
| Little impact | 17 | (23) | 8 | (12) |
| Moderate impact | 15 | (20) | 12 | (19) |
| Severe impact | 17 | (23) | 44 | (68) |
| Coping mechanism (%) |  |  |  |  |
| Current household income | 33 | (45) | 39 | (60) |
| Using savings | 55 | (74) | 9 | (14) |
| Asking relatives, religious bodies, non-government organizations | 34 | (46) | 47 | (72) |
| Borrowing from a financial institution | 3 | (4) | 6 | (9) |
| Cutting down on food and other purchases | 2 | (3) | 45 | (69) |
| Selling items (land, property, livestock, jewelry) | 4 | (5) | 13 | (20) |
| Withdrawing children from school | 0 | (0) | 3 | (5) |
| Reducing the number of medical visits | 0 | (0) | 1 | (2) |
| Other | 11 | (15) | 1 | (2) |
| Experiencing catastrophic costs, at threshold of (%) |  |  |  |  |
| 10% of household income | 63 | (85) | 58 | (89) |
| 15% of household income | 57 | (77) | 54 | (83) |
| 20% of household income | 48 | (65) | 49 | (75) |
| 25% of household income | 47 | (64) | 43 | (66) |
| 30% of household income | 44 | (60) | 41 | (63) |

\*Among participants with income-earning employment before cervical cancer diagnosis

The costs incurred for accessing CC-related services were mostly catastrophic for both countries. Using a threshold of 10% of annual household income, 85% of affected households in Bhutan and 89% of affected households in Zambia faced catastrophic costs. These proportions both decreased when we applied higher thresholds.

## Discussion

This study highlights the significant financial burden – notably high out-of-pocket expenses – faced by most women with CC and their households in Bhutan and Zambia. Expenses were catastrophic for over 85% of affected households in both countries. Participants reported severe financial impact, due to high DMCs, DNMCs, and ICs, particularly during the treatment phases. From a broader economic perspective, such burdens may hinder productivity and development in LMICs.

Using the COEUS framework, we applied two methodological approaches, to assess the CC-related financial burden in Bhutan and Zambia from a societal perspective. The different approaches also provided valuable insights into cross-country variations. The community outreach survey in Bhutan revealed diverse cost distributions across CC stages, while the facility-based survey in Zambia showed that the costs increased by the advanced stage. Differences observed are likely due to sampling. The facility-based survey probably captured participants adhering to treatment and living near hospitals, while the outreach surveys included harder-to-reach populations, including those not adhering to treatment. This may explain our unexpected finding of higher reported costs for stage 2 compared to stages 3 or 4 in Bhutan. It may be explained by the fact that individuals living far from hospitals are less likely to visit healthcare facilities and are diagnosed at more advanced stages. Women with stage 3 or 4 CC in Bhutan, in turn, often need treatment abroad in Thailand or India, which could negatively impact treatment compliance and result in lower reported costs.

In Bhutan, professional healthcare expenses were often accompanied by costs for alternative or traditional remedies including rituals, which are culturally important when faced with illness and death.[19] These alternative or traditional remedies – though not always affordable – were prioritized for their perceived spiritual and preventive value. The remedies costs varied greatly, from modest herbs and ceremonies to elaborate events, contributing to the wide variation in reported spending on non-professional care. Such practices underscore the need to consider cultural dimensions when evaluating financial burden and designing supportive interventions.

The high costs, assessed from a societal perspective, highlight the need for support beyond universal health coverage. In Bhutan, healthcare fees are covered by the government.[20] However, CC-affected households still pay for medicines that are not on an essential drug list as well as for private healthcare, both locally and abroad.[19] Women with CC received a daily allowance for themselves and their accompanying person when traveling abroad, of approximately Nu125 per day. Day or casual workers could also receive Nu15,000 to compensate for lost income. Additionally, many households of women with CC incur private accommodation expenses when public hospital wards are noisy or lack privacy.

Despite existing financial protection mechanisms, a substantial proportion of households continued to experience catastrophic costs. To cope with this burden, affected households could not only rely on their savings but also had to seek help from relatives or religious bodies. This reliance on social support networks is common practice and a social construct in LMICs, where extended household members often provide financial assistance with no expectation of repayment.[21, 22] Individuals who reported irregular and low incomes, may also possess assets that they can be sold to cope with unforeseen financial burdens.[23, 24] They may have engaged in the exchange of goods and services instead of money, as described by a woman with CC in this study. However, capturing all of these support mechanisms is challenging, as it is challenging to quantify the financial assistance received from relatives or the non-monetary exchange in goods and services.[25] This difficulty may lead to an overestimation of the proportion of households facing catastrophic costs.

This study has several limitations. First, the use of two distinct methodological approaches limit direct a comparison between the two countries, adding complexity to the interpretation of our results. Second, a substantial proportion of participants did not report their income, potentially leading to an underestimation of catastrophic costs. The main challenge in assessing the catastrophic financial burden in LMICs is accurately calculating household income, particularly among participants working in agricultural sectors such as farming, and where women may lack access to financial information due to cultural traditions that designate men as primary managers of household finances.

Despite the limitations in measuring catastrophic spending, the use of comment boxes in this study helped to elucidate our findings by complementing quantitative cost data with qualitative explanations. From this qualitative data, we realised that DMC, DNMCs, and ICs, were difficult to evaluate in LMICs like Bhutan and Zambia, and that they were potential underestimated. An alternative approach to assessing the catastrophic financial burden would have been to compare CC-related expenses to non-food essential household expenditures.[26] However, this approach would have required longer interview durations and may have potentially compromised the quality of the data we collected.

We provide one of the first comprehensive reports on the financial burden of CC in LMICs. Our findings highlight the importance of developing strategies to prevent and mitigate the catastrophic CC-related costs. The financial burden caused by CC, and its impact on families, hinders a country’s development. The burden can even result in children dropping out of school and teenage daughters being forced into marriage.[27] The global CC burden is projected to continue rising,[2] and will reach 700,000 cases and 400,000 deaths in 2030. As the burden rises, the financial and economic impact of this disease will also increase. Therefore, strategies for CC prevention through human papillomavirus vaccination and screening are crucial to counteract further increases in financial and economic burdens in LMICs.

## Conclusions

This study provides clear evidence that CC care seeking and treatment costs impose severe financial hardship on women and their households in Bhutan and Zambia. Expanding human papillomavirus vaccination, screening programs, and early detection efforts are essential to lower both the CC burden and the risk of catastrophic costs associated with this highly preventable disease.

## List of abbreviations

CC: cervical cancer
CDH: Cancer Disease Hospital COEUS (Costing Surveys to Assess the Economic Burden of Cervical Cancer in Society)
DMC: Direct Medical Cost
DNA: Deoxyribonucleic acid
DNMC: Direct Non-Medical Cost
HPV: Human Papillomavirus
IC: Indirect Cost
LEEP: Loop Electrosurgical Excision Procedure
LMICs: Low- and Middle-Income Countries
NGO: Non-Government Organization
Nu: Bhutan’s Ngultrum (currency)
WHO: World Health Organization
ZMK: Zambian Kwacha (currency)

## Declarations

### Ethics approval and consent to participate

We received ethical approval from the research ethics committees of the International Agency for Research in Cancer (IARC/WHO) (Ref No: IEC 22-21), the Ministry of Health of Bhutan (Ref No: REBH/Approval/2022/025), the University of Zambia Biomedical Research Ethics Committee (UNZABREC), and the National Health Research Authority of Zambia (Ref No: NHRA000015/8/12/2022) prior to study implementation. All participants received an explanation of the study and provided written consent before the interview. All data were pseudo-anonymized prior to analysis.

### Consent for publication

Not applicable.

### Availability of data and materials

Data are available upon request to the corresponding author. The request should be specific and will be assessed on a case-by-case basis by all authors. There are no personalised data in this study, but all data sharing will still abide by rules and policies defined by the involved parties. Data sharing mechanisms will ensure that the rights and privacy of individuals participating in research will be always protected.

## Authors’ contributions

AF conceived the original idea for the study and led the protocol and instrument development, data cleaning and validation analysis, and article writing. Pp, CK and NN reviewed the instruments, led data collection, and reviewed the report. VT and EL support the data collection and management. AF drafted the manuscript. DS, IM, IB. Pp, CK, and NN reviewed and edited the manuscript. IB and IM reviewed the analysis and results interpretation. IB supervised the project. All authors had full access to the data obtained from searching and extracted data and had the final responsibility for the decision to submit for publication.

## Disclaimers

Where authors are identified as personnel of the International Agency for Research on Cancer/World Health Organization, the authors alone are responsible for the views expressed in this article and they do not necessarily represent the decisions, policy, or views of the International Agency for Research on Cancer /World Health Organization. The designations used and the presentation of the material in this Article do not imply the expression of any opinion whatsoever on the part of WHO and the IARC about the legal status of any country, territory, city, or area, or of its authorities, or concerning the delimitation of its frontiers or boundaries. The findings and conclusions contained within are those of the authors and do not necessarily reflect positions or policies of the Gates Foundation.

## Acknowledgements

The authors thank Rachel Wittenauer (IARC/WHO) for suggestions to the manuscript and Nadia Akel (IARC/WHO) for English language editing support.

